# Capturing Social Prescribing in Primary Care Health Records: A Study of Coding Practices in England

**DOI:** 10.64898/2026.09.11.26362851

**Authors:** Feifei Bu, Daisy Fancourt

## Abstract

**Background:** Social prescribing has expanded rapidly in England and internationally. However, the collection of consistent data to monitor and evaluate its reach and impact remains to be challenging.

**Methods:** This study used routine data from the Clinical Practice Research Datalink (CPRD) Aurum in England. UpSet plots were used to describe intersections among different social prescribing codes, and between social prescribing and other non-clinical interventions. After identifying potential problematic intersections, we fitted multilevel regression models to examine variation between general practices and to understand factors associated with potentially problematic coding.

**Findings:** Between 2019 and 2025, approximately 3.8 million consultations involving social prescribing codes were recorded in CPRD Aurum. Social prescribing codes were most often recorded individually (38.1%), but intersections existed both among different social prescribing codes, and between social prescribing and related non-clinical support. Many intersections reflect plausible stages and scenarios within the social prescribing pathway. However, some coding patterns appeared ambiguous or potentially inconsistent, particularly those related to social prescribing being offered and referrals being made. Results from the multilevel regression models suggested that 28.4-34.6% of variation in these codes was attributable to differences between practices, and problematic coding varied by region and year of recording.

**Interpretation:** Our findings highlight opportunities to improve the quality and consistency of social prescribing coding in primary care, thereby strengthening service monitoring and evaluation both in England and internationally. They also provide important insights for researchers using CPRD data to investigate social prescribing.

## Introduction

Social prescribing (SP) is a mechanism of care which connects individuals to non-clinical support within their local communities tailored to their specific needs, interests and circumstances to improve their health and wellbeing [1]. As such, it represents a person-centred approach that is also co-productive in nature, typically involving a trusted individual, such as a social prescribing linker worker, who works collaboratively with the service user throughout the process [2]. SP reflects the recognition of the importance of social determinants of health in shaping individuals’ health outcomes and health inequalities across population groups, as well as the complex social, emotional and practical needs that are often intertwined with clinical conditions but not addressed by conventional medical care.

The origin of SP can be traced back to the Peckham Experiment began in 1920s [3]. And the Bromley by Bow Centre in London established in 1980s is considered as one of the earliest modern SP schemes [4,5], followed by a series of pilot projects across England and devolved nations in 1990s-2010s, such as the Arts for Well-being scheme in Durham [6], and the Rotherham Social Prescribing Pilot [7]. However, it was not until 2019, with the publication of the NHS Long Term Plan, that was SP formally established as a nationwide programme within NHS, as a key component of the Universal Personalised Care [8]. In this plan, NHS England committed to fund 1,000 link workers by 2020/21, with a target of 900,000 patient referrals by 2023/24. By 2022, SP became a formal mandate for every Primary Care Network (PCN) as part of its service provision, with funding available through the Additional Roles Reimbursement Scheme (ARRS) [9]. This commitment was further strengthened in the NHS Long Term Workforce Plan (2023), setting out a rising target of 9,000 link workers by 2036/37 [10].

With the national rollout of SP, it has become increasingly important to collect data to monitor its scale, reach and impact. In 2023, NHS England commissioned the Professional Record Standards Body (PRSB) to develop an information standard to support consistent recording of SP activities. Integral to this standard is a minimum dataset, which mandates the capture of patient demographics, needs and concerns, support offered, and outcomes. This requirement has been reiterated in subsequent documents, notably NHS England’s reference guide for PCNs [11]. However, capturing SP activities within the primary care systems remains challenging. Previous work has identified a comprehensive, although likely inexhaustive, list of SNOMED codes related to SP [12]. However, there remains ambiguity in how these codes are used in practice. For instance, it is unclear if the code “referral to social prescribing service” is used exclusively to denote completed referrals as intended [9], or if “social prescribing offered” may also be used to capture completed referrals.

In addition, there are several non-clinical support roles or programmes that exist alongside SP, some of which overlap or are used interchangeably with the link worker role. Examples include community navigator, community connector, wellbeing advisor that are used in local SP schemes before and even after moving toward a more standardised terminology from 2019 onwards [13,14]. However, other roles tend to have distinct origins and emphases. For example, health coach is generally framed as a supported self-care and self-management intervention for patients living with long-term conditions, which can be delivered by both clinicians and non-clinicians [15,16]. In contrast, health trainers, introduced in England through the 2004 public health White Paper Choosing Health [17], are typically recruited from local communities and trained to delivered behavioural changes interventions, with the aim of helping individuals adopt healthier lifestyles and reducing health inequalities, particularly in disadvantaged communities [18]. More recently, health and wellbeing coach, a dedicated non-clinical role, has become part of the NHS personalised care workforce in primary care and, like SP link worker, is funded through ARRS [19]. However, unlike SP link workers whose role centre on connecting people with community assets, health and wellbeing coaches focus more on internal capacity for change, including motivation, self-management and lifestyle change. While these roles are different to SP link workers, given the potential complementarity and overlap between them, it is important to delineate how they are used in real-world practice in relation to SP.

Therefore, the present study is set out to examine how SP is recorded within primary care systems, drawing on routine data from the Clinical Practice Research Datalink (CPRD). Our research questions were: (i) how are SP codes currently used, including which codes are most common, are they regularly used in isolation or in combination, and who typically enter these codes (RQ1); (ii) are there problematic patterns of code use (i.e. combinations of code that do not make logical sense) and how do they vary over time and geographically (RQ2); and (iii) how does use of SP codes intersect with codes for other non-clinical support roles or programmes that exist alongside SP (RQ3)? By systematically analysing the coding practice, this study seeks to identify the range, consistency, and practical use of relevant codes, as well as areas of ambiguity and variation in coding practices. In doing so, it contributes to a better understanding of the extent to which routine primary care data can reliably capture SP activities, with implications for service monitoring, evaluation and future data standardisation efforts.

## Methods

### Data

Data were from CPRD, a research data service that collects anonymised patient data routinely from a network of over 2000 general practices across the UK. CPRD contains rich data on demographics, diagnoses and symptoms, prescriptions, tests and referrals from over 60 million patients over more than 35 years since 1989, including over 18 million currently registered patients. This study focused on England using data from CPRD Aurum which covers over 20% of general practices and has been shown to be representative of the English population in terms of geographic area, deprivation, urbanicity, age and sex [20,21].

Using the December 2025 data build, we extracted all records matching any of the SP or related codes, as detailed in Table S1 in the supplementary material. Data extraction was conducted in January 2026 by an independent data analyst. In the analyses, we excluded records before 2019 to restrict the study period to the formal adoption of SP as a national programme. Further, we excluded records with missing consultation identifiers (∼6.8%) from the analyses.

### Measurement

For RQ1, the main outcome of interest was SP codes (11 codes including “social prescribing offered”, “referral to social prescribing service”, “social prescribing declined” etc.; Table S1).

For RQ2, problematic coding was defined as unclear use of offered code (i.e. a code indicating SP was offered without any information on whether the patient took up the offer), and inconsistent use of referral code (e.g. a code indicating a referral was made in combination with another code indicating it was not). A full definition of problematic codes is shown in Table 1. The main predictors included calendar year when SP and related activities were recorded (2019-2025), and geographical region of practices (North East, North West, Yorkshire & Humber, East Midlands, West Midlands, East of England, London, South East, South West).

**Table 1.** Definition of unclear or inconsistent use of offered and referral codes and intersection examples.

| Outcome | Code intersection examples (frequency>1,000) |
| --- | --- |
| Unclear use of offered code: offered coded used on its own or in combination with other codes but none of the following:<br>Referral to social prescribing service,<br>Referral to social prescribing service from other agency,<br>Social prescribing declined,<br>Not suitable for social prescribing | Social prescribing offered<br>Social prescribing offered +<br>Seen by social prescribing link worker<br>Social prescribing offered +<br>Social prescribing case closed<br>Social prescribing offered +<br>Social prescribing plan completed<br>Social prescribing offered +<br>Review of social prescribing plan<br>Social prescribing offered +<br>Seen by social prescribing link worker +<br>Social prescribing case closed<br>Social prescribing offered +<br>Review of social prescribing plan+<br>Social prescribing plan completed<br>Social prescribing offered +<br>Seen by social prescribing link worker +<br>Review of social prescribing plan<br>Social prescribing offered +<br>Social prescribing for mental health<br>Social prescribing offered +<br>Signposting to social prescribing<br>Social prescribing offered +<br>Social prescribing case closed +<br>Social prescribing plan completed |
| Inconsistent use of referral code: referral coded used in combination with “social prescribing declined” or “signposting to social prescribing” | Referral to social prescribing service +<br>Social prescribing declined<br>Referral to social prescribing service +<br>Signposting to social prescribing<br>Referral to social prescribing service +<br>Social prescribing offered+<br>Social prescribing declined<br>Referral to social prescribing service +<br>Social prescribing case closed +<br>Social prescribing declined<br>Referral to social prescribing service +<br>Seen by social prescribing link worker +<br>Social prescribing declined |

For RQ3, potentially related codes included roles such as community navigator, health coach, health trainer, health and wellbeing coach or worker (Table S1).

### Statistical analysis

Data were pooled across the full study period (2019-2025) and mainly analysed at consultation level based on consultation identifiers. Consultation was defined in a broad sense, including contacts with either clinical or non-clinical staff members. Results were presented descriptively in UpSet plots, a novel visualisation technique for quantitative analysis of intersecting sets and aggregates of intersections [22]. We first examined intersections between different SP codes (RQ1), and then assessed intersections between SP codes as a group and other related code groups (RQ3). Based on the descriptive analyses, we identified specific intersections requiring further investigation as outlined above (RQ2). We first fitted unconditional multilevel models to estimate variation across general practices, followed by conditional multilevel models including year as a predictor to examine changes over time, and region to access regional variations.

## Results

### RQ1: Intersections between SP codes and data coder

We explored the distribution of most common intersections (top 40 out of 477) among different SP codes across approximately 3.8 million consultations (Figure 1). In most consultations (38.1%), SP codes were used individually, although combinations of different codes were also observed. The most common intersection was between “social prescribing offered” and “referral to social prescribing service” (4.8%), followed by the intersection between “referral to social prescribing service” and “seen by social prescribing link worker” (1.6%).

**Figure 1.**
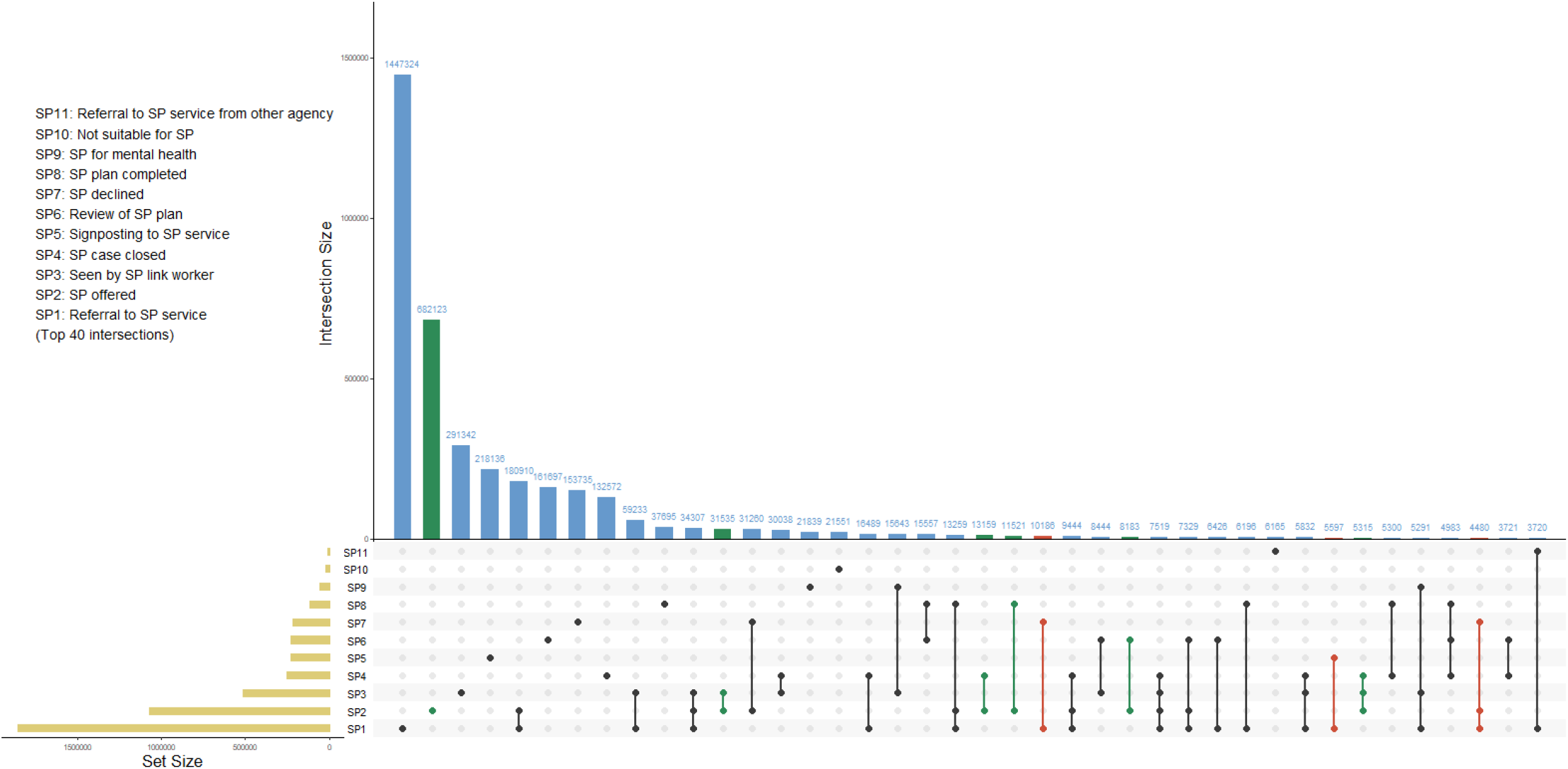
Intersections between different social prescribing codes in CPRD

Overall, the observed intersections were largely coherent as expected. However, some intersections appeared potentially inconsistent or difficult to interpret. One notable issue concerned the use of “social prescribing offered”. When recorded on its own (18.0%), the outcome of the offer remained unclear, without indicating whether the patient declined or were referred to SP. In addition, this code was used in combination with “seen by social prescribing link worker”, “social prescribing plan completed”, and “review of social prescribing plan”. Although these intersections were relatively rare, they suggested that the offered code could have been used interchangeably with “referral to social prescribing service”.

A second notable issue related to the code “referral to social prescribing service”. Some observed intersections appeared conceptually inconsistent, such as the combination of “referral to social prescribing service” and “social prescribing declined” within the same consultation (n=21,713). In 95.3% of cases (n=20,682), these two codes were entered by the same staff member on the same day, with only 1.7% by different people on the same day, and another 3.1% were entered on different days (n=668; nearly all by the same staff member). Another example was “referral to social prescribing service” in combination with “signposting to social prescribing service” (n=9,741). Among them, over 99.6% were recorded on the same day, but 24.3% were recorded by different staff members, which might, to some extent, explained the inconsistency.

SP data were entered by both clinical and non-clinical staff across a wide range of job titles, with the most common staff categories being general medical practitioner (12.3%), SP link worker (12.1%), health care support worker (9.7%) (Figure S1). Administrative and clerical staff also entered SP data relatively commonly. It is of note that the distribution of staff roles varied considerably across SP codes (Figure S2). For instance, the signposting code was largely recorded by managers and administrative staff. The “not suitable for social prescribing” code was mostly commonly recorded by nurses (20.6%), followed by general medical practitioners (17.7%) and pharmacists (12.4%).

### RQ2: Patterns of problematic code use

Offered code: As shown in Figure 2a, the percentage of consultations using the offered code in an unclear way was 27.5% in 2019 which decreased year on year, suggesting a gradual improvement over time. Nevertheless, it still accounted for 16.1% of all consultations in 2025, and the percentage of practices using the code unclearly remained high (88.9%; albeit down from a peak of 98.8% in 2023), suggesting that this was a widespread pattern across practices rather than driven by a small number of outliers.

**Figure 2.**
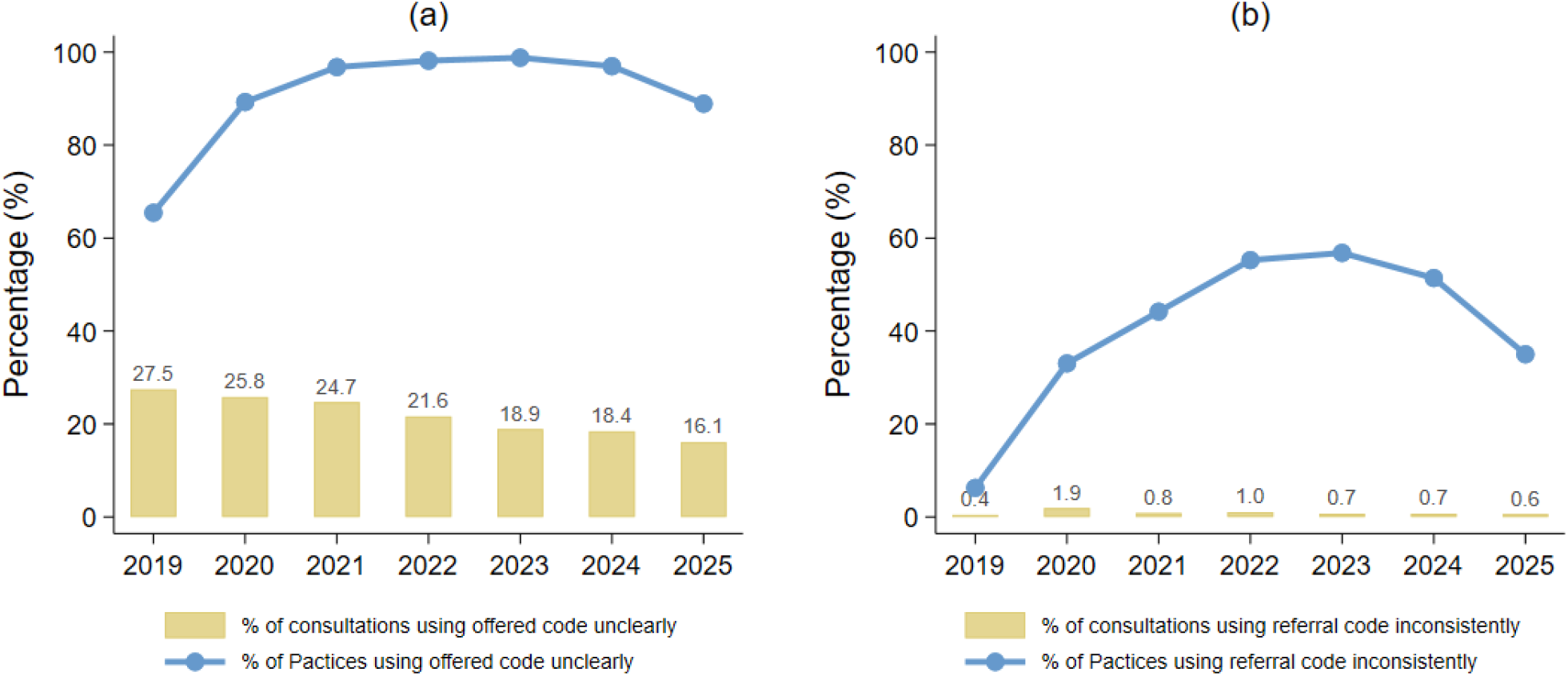
Trends in the unclear use of the offered and referral codes in CPRD (2019-2025)

Based on the unconditional regression model, approximately 28.4% of variation in the unclear use of offered code was attributable to differences between practices (Table 2). Results from the conditional model confirmed that unclear use of the code reduced over time. Moreover, practices in West Midlands and East of England regions were less likely to record the code unclearly compared to those in North East.

**Table 2.**
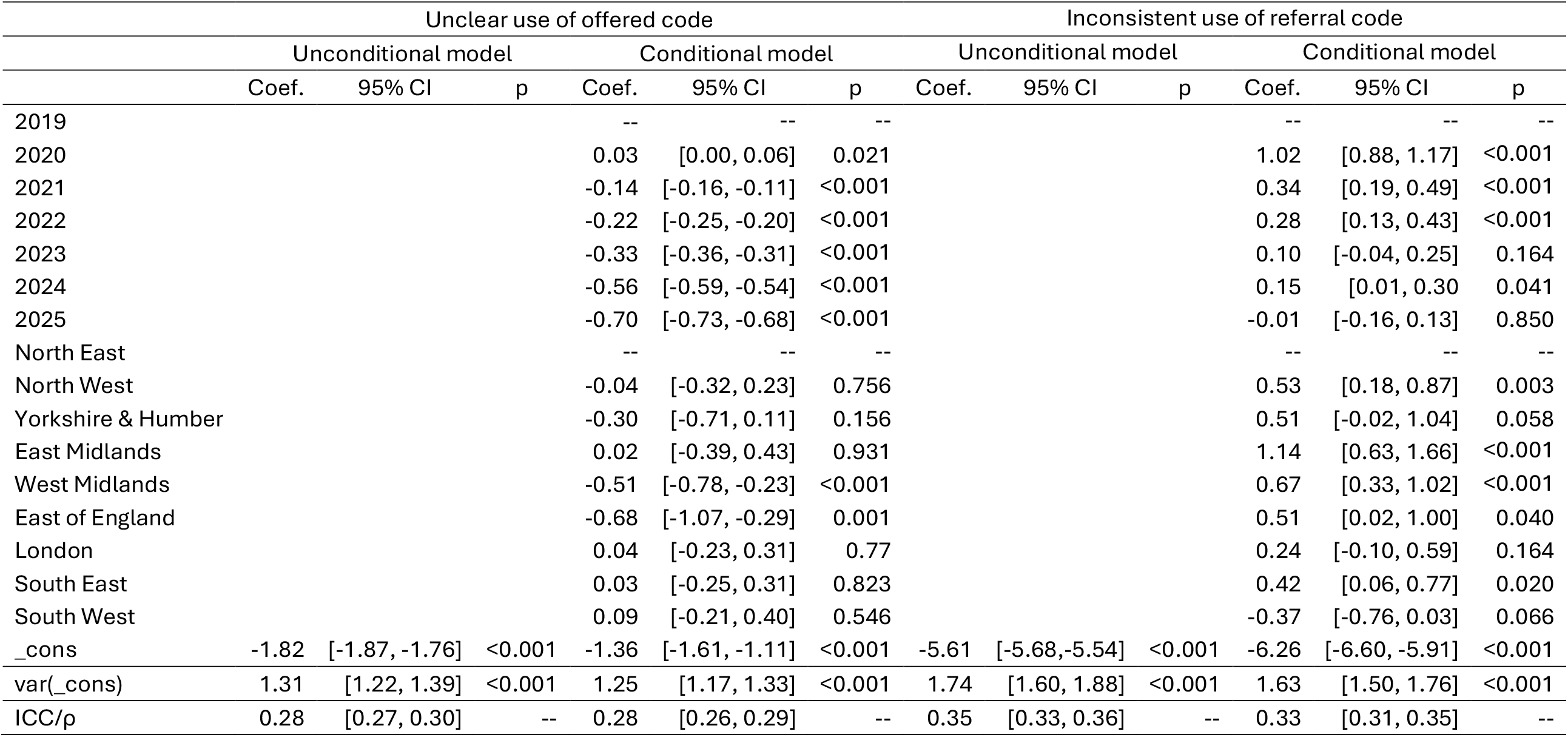
Results from multilevel mixed-effects logistic regression models.

Referral code: The percentage of consultations using the referral code inconsistently was generally very low (0.4-1.9%), with a slight improvement from 2020 onwards (Figure 2b). The percentage of practices using it inconsistently increased between 2019 and 2023, before starting to decrease. However, there were still 35.0% of practices recording the referral code inconsistently in 2025.

Results from multilevel regression models showed that around 34.6% of variation was attributable to between-practice differences (Table 2). The conditional model indicate that inconsistent use was more common from years 2020 to 2022 than 2019, with limited evidence for more recent years. Regional differences were also observed, with practices in the North West, East Midlands, West Midlands, East of England and South East more likely to record the referral code inconsistently.

### RQ3: Intersections of SP with other related codes

Overall, SP codes were predominantly used in isolation from other related codes such as community navigator, health and wellbeing coach or worker, health coach and health trainer (Figure 3). However, there was some evidence of combined usage of SP with other codes, accounting for 2.4% of cases when SP codes were recorded. The most common intersection was between SP and health and wellbeing coach or worker, followed by intersections with community navigator, health coach and health trainer codes.

**Figure 3.**
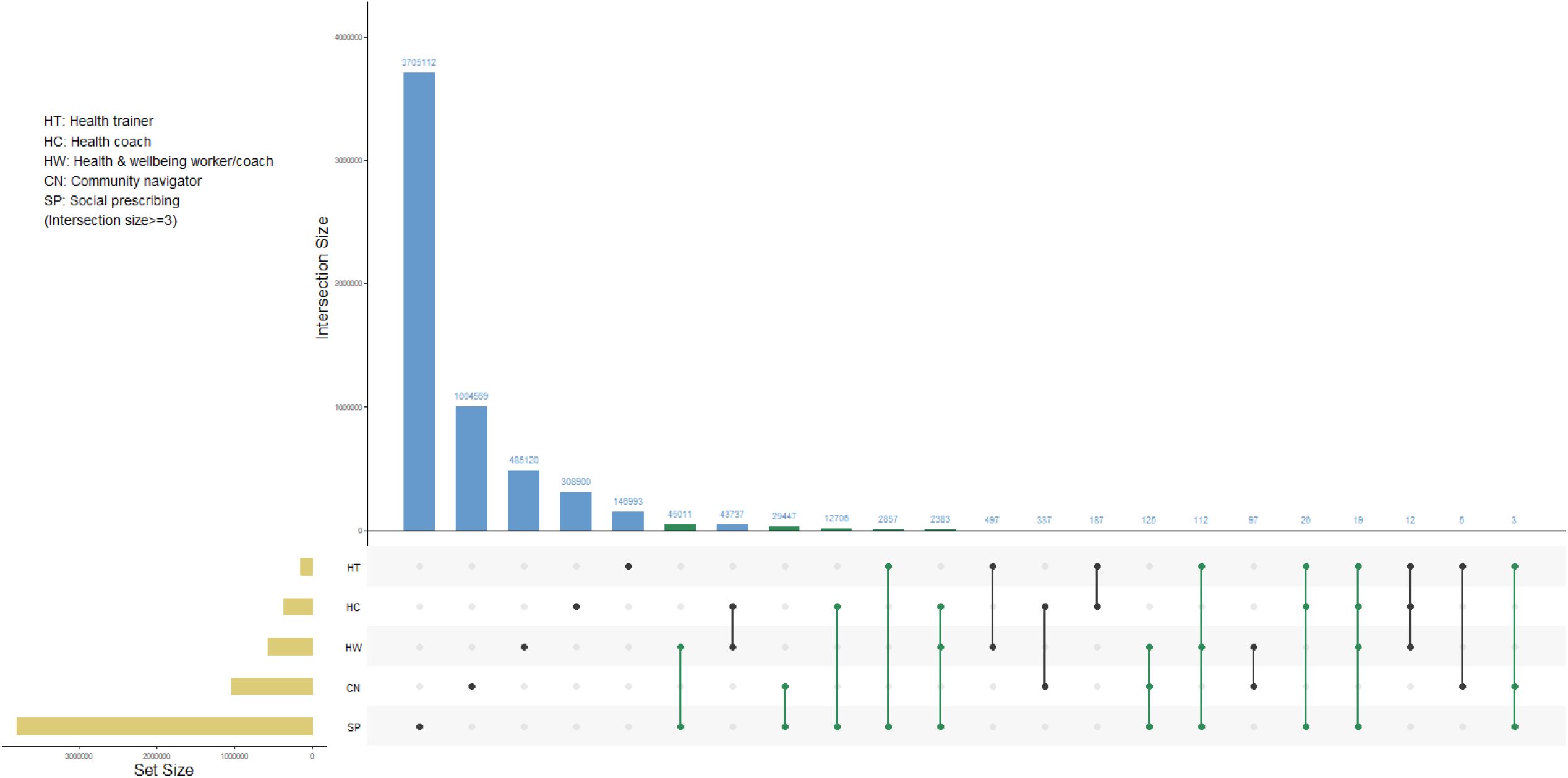
Intersection of social prescribing with other related codes in CPRD

Analyses examining these patterns in greater detail (moving beyond code groups to explore specific code intersections for each pair of code groups; Figures S3–S6) showed that the most common intersection was SP referral or offered codes recorded in combination with a “seen by” code, including health and wellbeing coach, community navigator and health coach. Health trainer was an exception, for which the most common intersection was between “referral to social prescribing service” and “referral to health trainer declined”, followed by “referral to social prescribing service” and “referral to health trainer”. It was notable that, for health trainer, some common intersections involved both “seen by health trainer” and “seen by social prescribing link worker”. Altogether, they accounted for 16.6% of all health trainer-SP intersections. In contrast, equivalent intersections accounted for only 2.2% for health and wellbeing coach, and less than 1% for health coach and community navigator.

## Discussion

This study examined how SP was recorded in routine primary care data following its national roll-out in England. Across approximately 3.8 million consultations from 2019-2025 in CPRD Aurum, the data suggest that SP codes are most often recorded individually, but intersections exist both across different SP codes, and between SP and related non-clinical support. Many intersections reflect plausible stages and scenarios within the SP pathway. However, some coding patterns are ambiguous or potentially inconsistent. Below, we discuss some of the most prominent issues and propose recommendations for researchers using these data, as well as policymakers and practitioners involved in service implementation, monitoring and evaluation.

The most prominent issue identified is related to the “social prescribing offered” code, particularly given that it is among the most frequently used code, second only to “referral to social prescribing service”. When recorded in isolation, this code does not indicate whether the offer is accepted or declined, but its intersections with other SP codes, such as “seen by social prescribing link worker”, “social prescribing plan completed”, “review of social prescribing plan”, suggest that it may sometimes be used in place of “referral to social prescribing service”. Although unclear use of this code has decreased over time, it remains common and was observed across the majority of practices in 2025, despite not being one of the NHS-recommended codes for recording SP activities in primary care [11]. The continued widespread use of this code has important implications for researchers using routine primary care data to study SP. In particular, relying exclusively on NHS-recommended codes is likely to underestimate SP activities to some extent. Accuracy may be improved by incorporating relevant code combinations that indicate acceptance, referral, link worker contact, or plan development at a later stage of SP pathway.

Another issue concerns the “referral to social prescribing” code. Although inconsistent use of this code is generally rare compared with the offered code, several intersections are worth noting. The most notable example is the combination of referral and decline codes, which is conceptually contradictory, as “referral to social prescribing service” is intended to indicate completed referrals [9]. Since most of these records were entered on the same day by the same staff member, this intersection is more likely to reflect uncertainty in coding rather than sequential changes in patient preference. Another ambiguous pattern is the combination of referral and signposting codes. Since signposting and referral imply different levels of support and engagement, their combined use raises questions about whether these codes are being used to capture distinct activities or are being applied inconsistently. Therefore, for researchers who are interested in referral, either as an outcome or exposure, additional steps may be needed to refine referral definitions by incorporating other SP codes, particularly the declined code.

The overlap between SP and related non-clinical support further complicates interpretation. Specifically, common intersections between SP referral or offered codes and the “seen by” codes for related non-clinical roles may indicate complementary care, where individuals are referred through SP and subsequently supported by another role. This is likely given non-clinical roles, such as health and wellbeing coach, are expected to work closely in partnership with SP link workers or service providers [23]. However, these intersections may also reflect role overlap or interchangeable terminology. This is especially likely for community navigators, which are in some settings described as equivalent to SP link workers [13,14]. For health and wellbeing coaches and health coaches, however, the interpretation is less clear. Routine primary care data alone cannot determine whether they represent distinct services, sequential support within a shared pathway, or local variation in terminology and role boundaries.

The distinct pattern observed for health trainer codes is also noteworthy. Health trainers historically focus on providing direct support for lifestyle change and reducing health inequalities [18], whereas SP link workers have a broader remit, primarily involving onward referral and connection to a wider range of community-based support [24]. The frequent intersection between “referral to social prescribing service” and “referral to health trainer declined” may indicate that SP are sometimes considered as an alternative when referral to health trainer is declined. However, they can also complement each other in some circumstances, as indicated by intersections between “seen by social prescribing link worker” and “seen by health trainer”. This pattern was distinctive to health trainer codes, suggesting health trainer is less likely to be used interchangeable with SP link worker compared to other non-clinical roles, namely community navigator, health and wellbeing coach and health coach.

Regarding the unclear or inconsistent use of SP codes, we found meaningful variation across practices and regions. For instance, multilevel analyses show that approximately 28.4% of variation in unclear use of the offered code, and around 34.6% of variation in inconsistent use of the referral code, was attributable to differences between practices. Regional patterns are also evident. For instance, practices in West Midlands and East of England are less likely to use the offered code in an unclear way, but more likely to use the referral code inconsistently. Taken together, these findings suggest that ambiguity in how SP is recorded is shaped not only by the codes themselves, but also by local coding conventions, staff training, service models and so forth. As such, we propose a series of recommendations to improve data capture and interpretation.

First, there is a need for clearer guidance on the use of the full set of codes directly related to SP. These codes appear to correspond to different stages of the SP pathway, including offer, referral, decline, contact with a link worker, care planning, review and completion. Guidance should therefore specify which codes should be used at which stage of the pathway, what each code is intended to capture, and which staff members are expected to record them. Second, the role of the SP offered code should be reconsidered. When used in combination with other codes, it provides limited additional information beyond codes indicating referral, decline, or link worker contact. When used in isolation, however, it is difficult to interpret because it does not indicate outcome of the offer. Despite not being one of the NHS-recommended codes, it is still widely used. We therefore recommend removing it from the system. Third, greater clarity is needed to define role boundaries between SP link workers and related non-clinical roles. This is crucial for understanding how different non-clinical roles operate within a holistic care model, and for distinguishing genuine complementary working from overlapping responsibilities or interchangeable terminology. Finally, to address the inherent limitations of primary care health records in capturing SP activities, including time pressures and system constraints, these records could be supplemented with data from specialised platforms such as Access Elemental and Joy. However, the full value of these complementary data sources can only be realised through effective data linkage.

## Conclusions

To the best of our knowledge, this is the first study that examines how SP and related non-clinical interventions are recorded in routine primary care data. It provides important implications for researchers, policymakers and practitioners, highlighting both the potential value of these data and the challenges involved in interpreting them. Further research is needed to understand the complexity of SP recording and service delivery. In parallel, policy action is needed to improve the quality and consistency of SP recording through standardised guidance, staff training and a simplified code list. Such improvements are crucial for accurate service monitoring, robust evaluation and the continued development of SP services.

## Article information

### Data availability

The original data from this study are provided by Clinical Practice Research Datalink (CPRD). Data cannot be shared publicly because they are not publicly available. CPRD may provide researchers with data following completion of their approvals and ethics process: https://www.cprd.com/research-applications

### Ethics statements

Ethical approval for this study was obtained from the Independent Scientific Advisory Committee of CPRD (protocol no. 25_005518).

### Patient and Public Involvement

Patients or the public were not involved in the design, or conduct, or reporting, or dissemination plans of our research

## Acknowledgement

This study is supported by the Economic and Social Research Council (ESRC) [UKRI1717], and the National Academy for Social Prescribing (NASP). We are grateful to colleagues at NASP for their helpful discussions and feedback.

