## Supplement for "Capturing Social Prescribing in Primary Care Health Records: A Study of Coding Practices in England"

### Supplementary Materials

Table S1. SNOMED code list for social prescribing and other potentially related activities

| Group | Term |
|---|---|
| Social prescribing | Referral to social prescribing service |
| | Social prescribing offered |
| | Seen by social prescribing link worker |
| | Social prescribing case closed |
| | Signposting to social prescribing service |
| | Review of social prescribing plan |
| | Social prescribing declined |
| | Social prescribing plan completed |
| | Social prescribing for mental health |
| | Not suitable for social prescribing |
| | Referral to social prescribing service from other agency |
| Community navigator | Seen by community navigator |
| | Referral to community navigator |
| Health coach | Health coach initiated encounter |
| | Referred for health coaching |
| | Seen by health coach |
| | Patient initiated health coach encounter |
| | Health coach |
| | Referral for health coaching |
| Health trainer | Referral to health trainer declined |
| | Referral to health trainer |
| | Seen by health trainer |
| | Signposting to health trainer |
| Health & wellbeing | Seen by health and wellbeing coach |
| | Signposting to health and wellbeing worker |
| | Health and wellbeing plan |
| | Health and wellbeing plan reviewed |
| | Health and wellbeing plan completed |

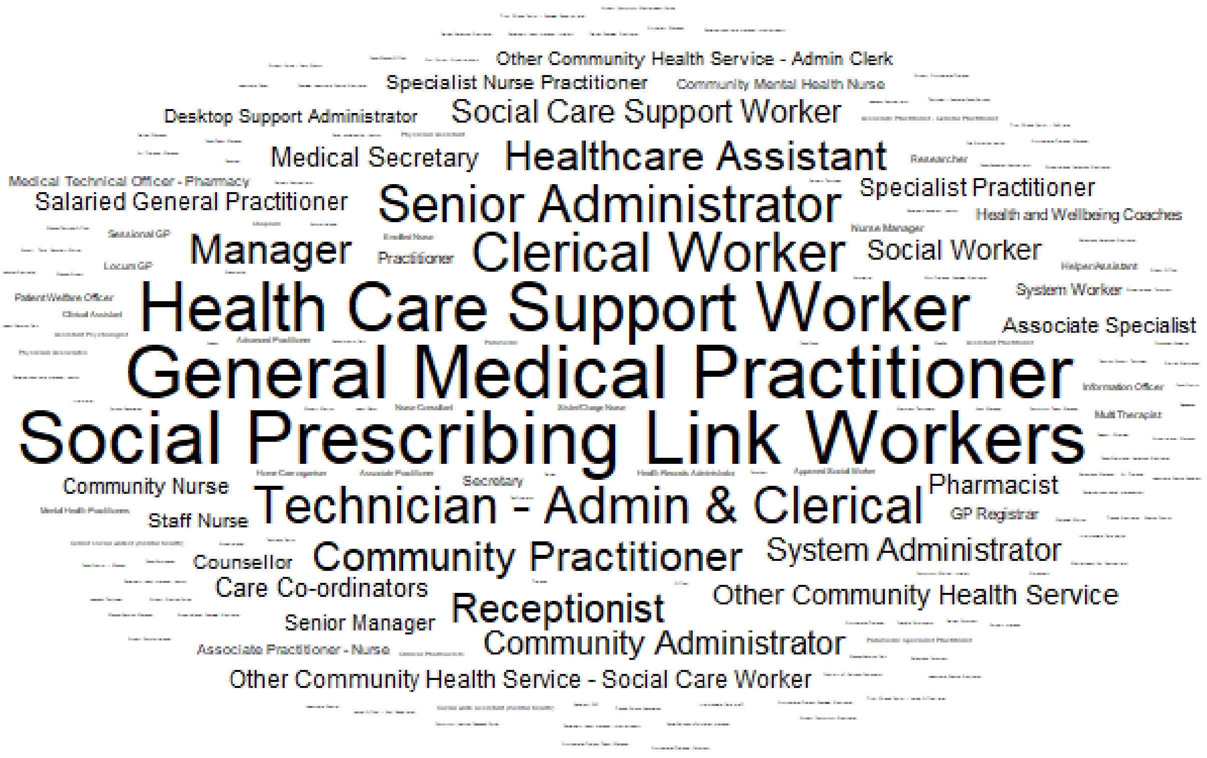


Figure S1 Job titles of staff members who recorded any social prescribing codes

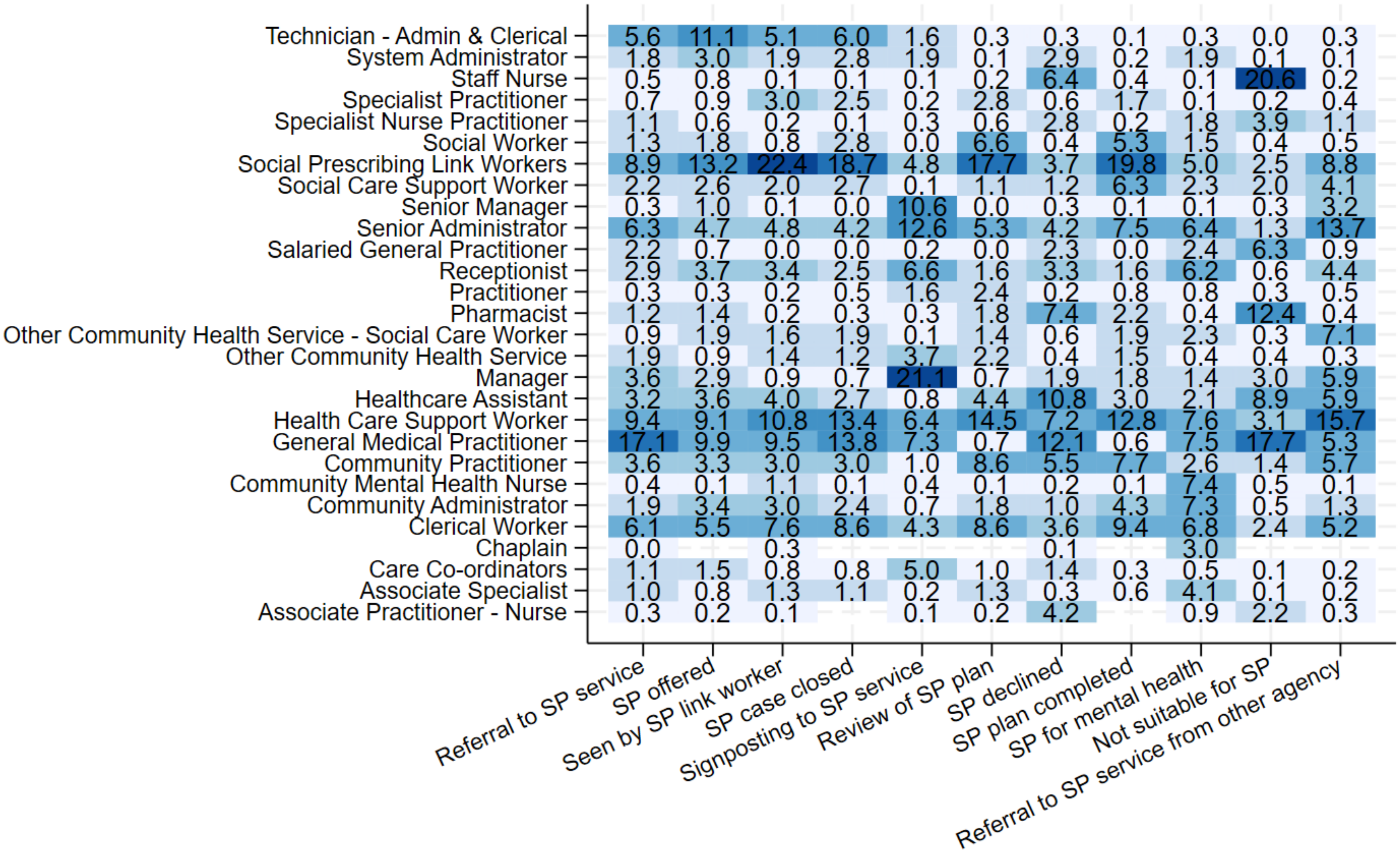


Figure S2. Job titles of staff members who recorded each specific social prescribing codes (including only top 10 job titles in any of the codes)

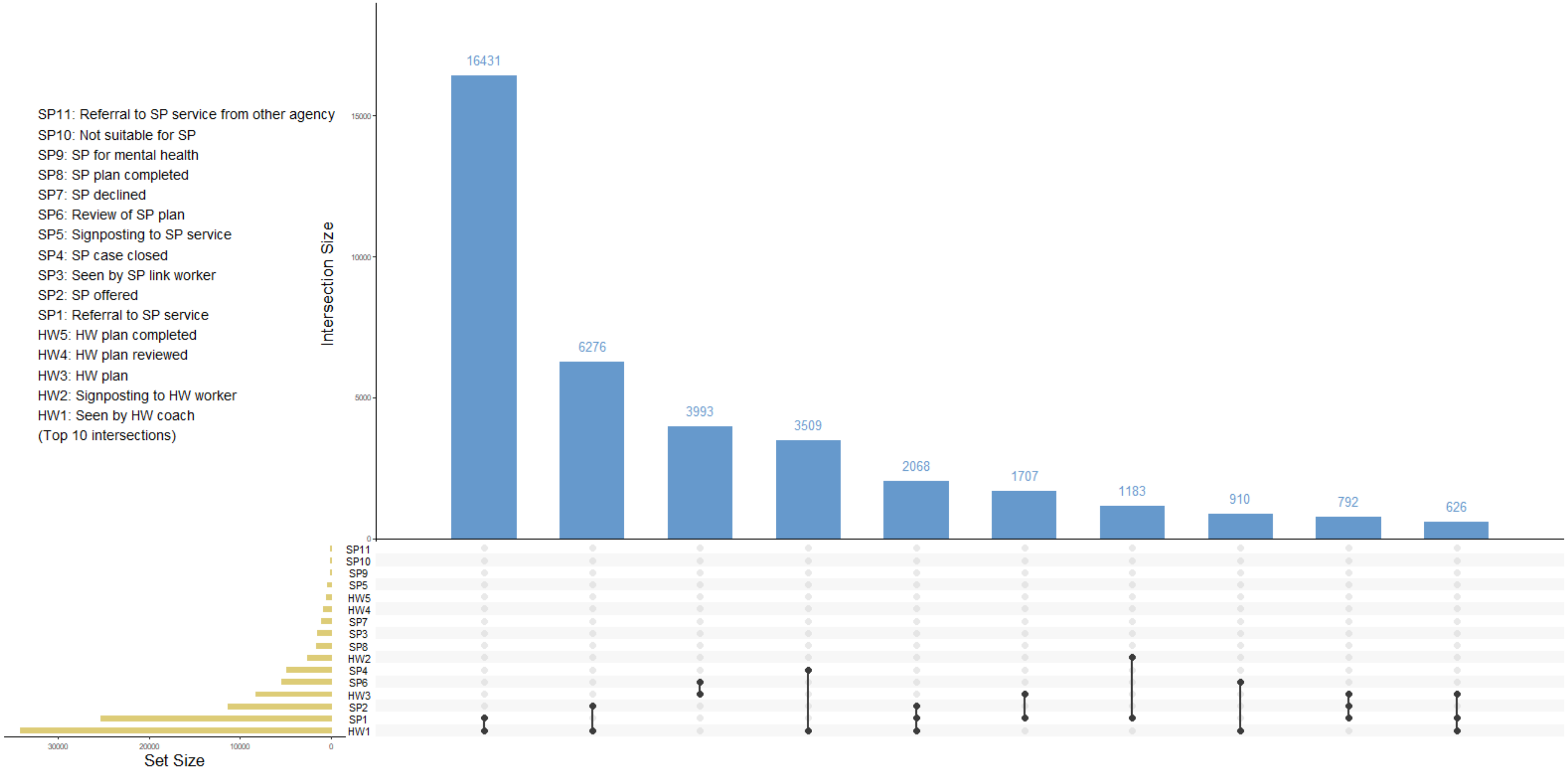


Figure S3. Intersections between social prescribing and health and wellbeing codes

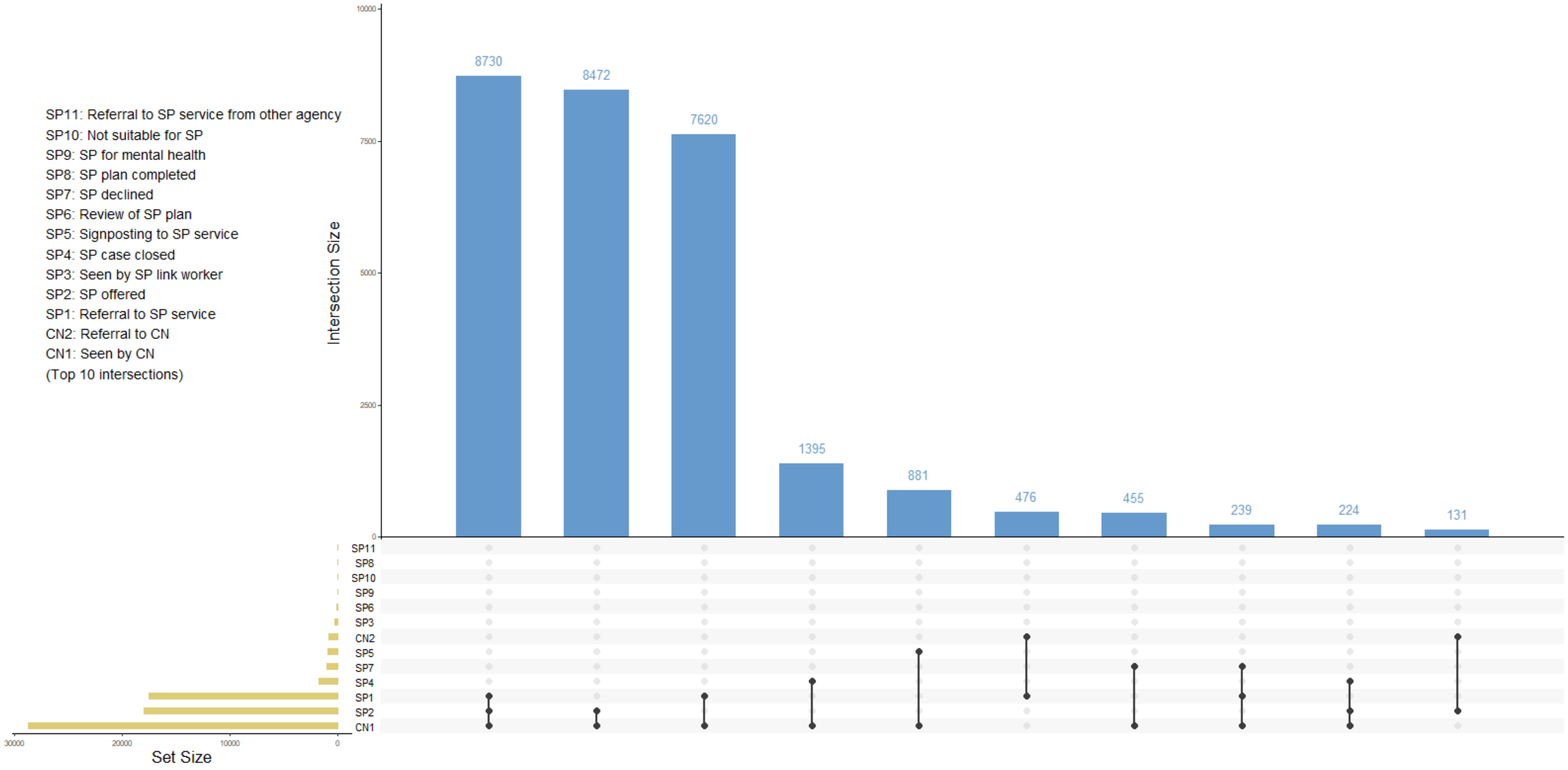


Figure S4. Intersections between social prescribing and community navigator codes

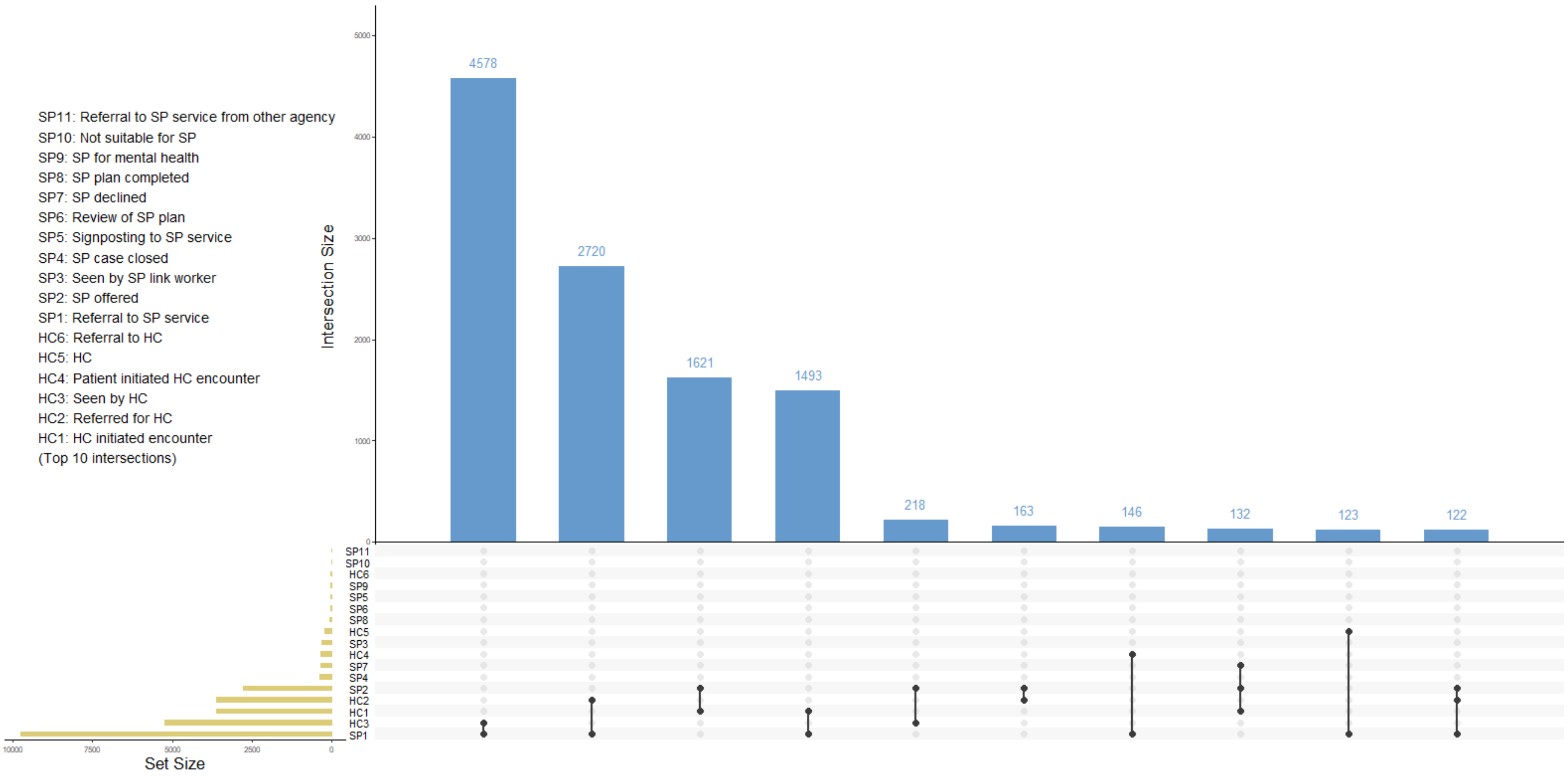


Figure S5. Intersections between social prescribing and health coach codes

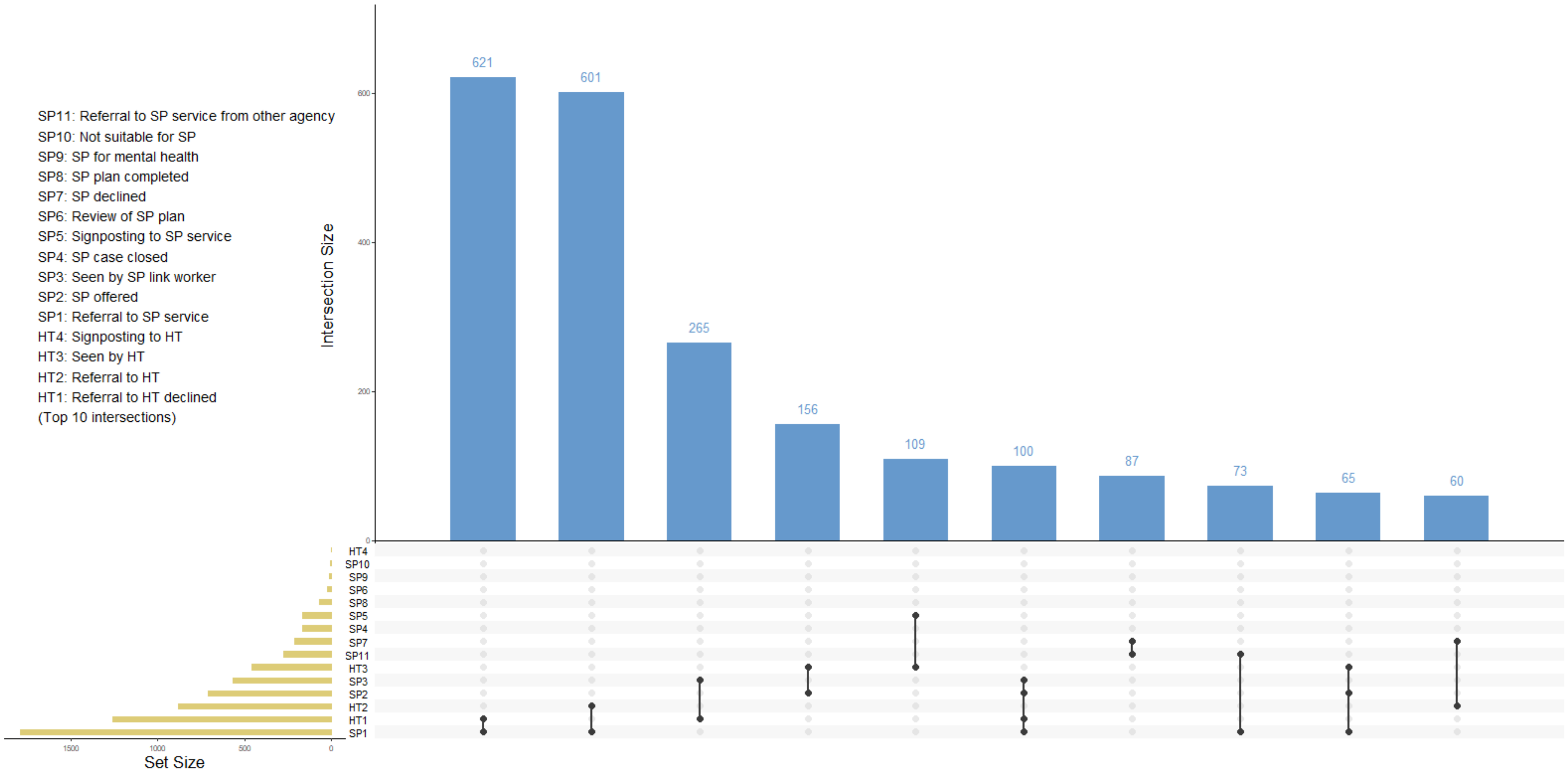


Figure S6. Intersections between social prescribing and health trainer codes
